# Epidemiology and Phylogenomic Characterization of Distinct 2023 and 2024 Mpox outbreaks in Kinshasa, Democratic Republic of the Congo - Evidence for increasingly sustained human-to-human transmission of subclade Ia

**DOI:** 10.1101/2024.11.15.24317404

**Authors:** Tony Wawina-Bokalanga, Sydney Merritt, Eddy Kinganda-Lusamaki, Daan Jansen, Megan Halbrook, Elisabeth Pukuta-Simbu, Áine O’Toole, Emmanuel Hasivirwe Vakaniaki, Rilia Ola-Mpumbe, Papy Kwete-Mbokama, Prince Akil-Bandali, Cris Kacita, Ange Ponga-Museme, Nelson Mapenzi-Kashali, Adrienne Amuri-Aziza, Olivier Tshiani-Mbaya, Princesse Paku-Tshambu, Pedro HLF Dantas, Tessa de Block, Emmanuel Lokilo, Chloé Muswamba, Jean-Claude Makangara-Cigolo, Gradi Luakanda, Christian Ngandu, David J. Kelvin, Catherine Pratt, Ahidjo Ayouba, Sofonias Tessema, Nicaise Ndembi, Antonio Mauro Rezende, Eric Delaporte, Dieudonné Mwamba, Lorenzo Subissi, Laurens Liesenborghs, Nicole A. Hoff, Martine Peeters, Nicola Low, Steve Ahuka-Mundeke, Jean-Jacques Muyembe-Tamfum, Anne W. Rimoin, Jason Kindrachuk, Koen Vercauteren, Andrew Rambaut, Placide Mbala-Kingebeni

**Affiliations:** Institut National de Recherche Biomédicale (INRB), Kinshasa, Democratic Republic of the Congo; Service de Microbiologie, Département de Biologie Médicale, Cliniques Universitaires de Kinshasa, Université de Kinshasa, Democratic Republic of the Congo; Department of Clinical Sciences, Institute of Tropical Medicine, Antwerp, Belgium; Department of Epidemiology, Jonathan and Karin Fielding School of Public Health, University of California, Los Angeles, CA, USA; TransVIHMI, Université de Montpellier, INSERM, IRD, Montpellier, France; Institute of Ecology and Evolution, University of Edinburgh, Edinburgh, UK; Institut National de Santé Publique (INSP), Kinshasa, Democratic Republic of the Congo; Graduate School of Cellular and Biomedical Sciences, University of Bern, Bern, Switzerland; Department of Microbiology and Immunology, Faculty of Medicine. Dalhousie University, Halifax, NS, Canada; Biosurv International, Salisbury SP4 0DQ, England; Africa Centers for Disease Control and Prevention (Africa CDC), Addis Ababa, Ethiopia; Instituto René Rachou, Fiocruz Minas, Belo Horizonte, Brazil; World Health Organization, Geneva, Switzerland; Department of Microbiology, Immunology and Transplantation, KU Leuven, Leuven, Belgium; Institute of Social and Preventive Medicine, University of Bern, Bern, Switzerland; Department of Medical Microbiology & Infectious Diseases, Max Rady College of Medicine, University of Manitoba, Winnipeg, MB, Canada

## Abstract

**Background:** Monkeypox virus (MPXV) is endemic to the Democratic Republic of the Congo (DRC), and historically has been reported in rural, forested regions with limited household and primarily zoonotic transmission. With the emergence of subclade Ib and increasing reported human-to-human transmission of MPXV we sought to characterize both the epidemiologic and genomic characteristics of confirmed mpox cases in Kinshasa, DRC, following confirmed MPXV introduction in August 2023.

**Methods:** This retrospective observational study leveraged demographic and case-investigation data, as well as biological samples, collected from suspected mpox cases in Kinshasa from January 2023 to September 2024 as part of routine national surveillance. Samples were tested at the Institut National de Recherche Biomédicale first by qPCR to confirm MPXV, and subsequently sequenced for genomic analysis for those samples with a cycle threshold value <31.

**Findings:** Epidemiological characterization of all confirmed mpox cases in Kinshasa indicated two distinct periods of MPXV introduction and circulation: first, following the initial case in August 2023, self-limiting transmission chains of subclade Ia primarily among men. Case investigations from this period suggest human-human transmission. The second period is marked by the introduction of subclade Ib to the city and expanded co-circulation of both subclades – with Limete health zone reporting the most cases of both subclades. Of those samples sequenced, genomic analysis indicated that greater than 60% of mutations in both subclades were consistent with APOBEC3-driven mutations.

**Interpretation:** These data suggest that as of July 2024, MPXV in Kinshasa has undergone an epidemiological shift with sustained human-to-human transmission and co-circulation of two distinct subclades. Additionally, high rates of APOBEC3-driven changes further support this shift towards human-to-human transmission. While sexual-contact transmission cannot be confirmed without further case investigation, these data highlight important changes in mpox dynamics which may better guide mpox mitigation and containment strategies in Kinshasa.

**RESEARCH IN CONTEXT:** 

**Evidence before this study:** Historically, human infections with Clade I monkeypox virus (MPXV) have been driven through zoonotic transmission with limited secondary transmission. However, sexual contact-mediated transmission of Clade I MPXV among a small cluster of cases was identified for the first time in Kwango Province, DRC, in mid-2023. We searched PubMed, bioRxiv, and medRxiv for articles published from database inception to 11 November 2024, with no language restrictions, using the terms “mpox, monkeypox, Congo Basin, Clade I, AND outbreak”. All articles were published from 2010, with the majority published from 2022-2024, and included reviews, case reports, and laboratory investigations. Early reports on mpox outbreaks demonstrated that most infections resulted from zoonotic transmission events and the risks of sustained human-to-human transmission were considered to be very low. Assessment of human-to-human transmission of MPXV in the DRC between 1970-1980 and 1981-1986 suggested that there was no evidence for changes in secondary attack rates. This included a study of 93 MPXV-infected individuals who became symptomatic following close contact with mpox patients from 1981-1986 in DRC. Here, 69 cases resulted from transmission during the first virus generation, 19 in the second generation, and five cases in both the third and fourth generation. During the 1996-1997 mpox outbreak in DRC, it was reported that 73% of cases had contact with an mpox patient. A study of Clade I mpox in the Republic of the Congo from April-July 2003 suggested that up to seven sequential virus generations had occurred during the outbreak and was supported by the clustering of cases, uniform intervals of cases, and prior exposure histories of the patients. High secondary attack rates were also reported during an outbreak of Clade I mpox from July-December 2013 in the DRC where transmissions within the community were estimated to be ≥7 virus generations. While these observations suggested potential increased patterns of human-to-human MPXV transmission, these continued to be linked to close, sustained contacts primarily within households and among caregivers. However, the first report of a cluster of mpox cases in DRC linked to sexual contacts in 2023 suggested that intimate contact, including through heterosexual and same-sex contacts, could result in Clade I MPXV transmission. Viral genome sequences isolated from this case cluster demonstrated the highest sequence similarity with a 2022 Clade I MPXV sequence from DRC. Subsequent investigation of surveillance data in South Kivu province, DRC, from September 2023 to January 2024 identified 241 suspected mpox cases that included sustained human-to-human transmission. Among PCR-confirmed mpox cases, the median age was 22 years with 29% of individuals identifying as sex workers. Genomic analysis identified a distinct clade I lineage, named as subclade Ib, with a predominance of APOBEC3 mutations. Rapid geographic expansion of subclade Ib has resulted in introduction of MPXV to multiple neighboring countries for the first time and including sustained chains of virus transmission, most notably in Burundi. Subclade Ia MPXV has undergone rapid and extensive geographic expansion in DRC since 2023. A recent longitudinal analysis of MPXV genomes from 2018-2024 demonstrated that all non-South Kivu Province sequences exhibited high genetic diversity, low incidences of APOBEC3 mutations as compared to subclade Ib, and likely resulted from multiple zoonotic introductions. Collectively, historic mpox case data and recent observations since 2023 have suggested that human mpox cases in DRC have been driven by zoonotic transmissions of subclade Ia and an increasing burden of sustained human-to-human transmissions of subclade Ib in Eastern DRC, with limited disease burden in Kinshasa.

**Added value of this study:** This investigation demonstrates a shift in the epidemiology of subclade Ia group II MPXV outbreaks, which had previously been typified by multiple independent zoonotic introductions. Epidemiologic analysis indicated two distinct periods for the Kinshasa mpox outbreak - an initial period from August 2023 to June 2024 characterized by sporadic subclade Ia mpox case detections, and a second period from July 2024-October 2024 characterized by co-emergence of both subclade Ia and Ib. The majority of mpox cases were adult males. In contrast to historic observations, children <15 years were not the primary demographic overrepresented among the subclade Ia mpox cases with those aged 16–49 years accounting for 79% of cases in this study. This investigation sequenced samples from mpox-confirmed cases in Kinshasa, DRC, by generating 115 near-complete MPXV genomes accompanied by phylogenomic analysis. Our findings reveal an escalation of the Kinshasa mpox outbreak that includes the establishment of both subclade Ia (Group II) and subclade Ib MPXV. Subclade Ia introductions (n=11 samples belonging to 5 clusters within Group II) into Kinshasa were observed in 2023 and 2024 with limited local transmission chains. In this analysis, we estimate that 63% of mutations in subclade Ia and 66% of mutations in subclade Ib from MPXV genomes collected in Kinshasa were consistent with APOBEC3-driven changes, a hallmark of human-to-human transmission.

**Implications of all the available evidence:** This study provides critical evidence for a rapid shift in subclade Ia MPXV epidemiology to include more sustained human-to-human transmission. While Clade I mpox cases have historically been linked to zoonotic transmissions, the recent emergence of subclade Ib in Eastern DRC coupled with our observations reported here for subclade Ia further complicate our understanding of Clade I MPXV transmission. These results have important public health implications for regional and international partners given Kinshasa’s role as an international travel hub and as the largest city on the continent. Observations of new sequences from Kwilu, Kwango, and Kongo-Central provinces (subclade Ia), as well as Kasaï, Tanganyika and Tshopo (subclade Ib), linked to Kinshasa genomes further increase concerns regarding the broader risks for subclade Ia expansion, including within sexual networks.

## INTRODUCTION

Monkeypox virus (MPXV), the etiologic agent of mpox, is phylogenetically classified into two major clades: Clade I and Clade II [1]. Both virus clades have now been subdivided, following the identification of genetically distinct variants in 2022 and 2024. Clade I includes subclades Ia and Ib, and Clade II, subclades IIa and IIb [2, 3]. In association with evolving phylogeny, the epidemiology of MPXV transmission has also continued to evolve, following re-emergence of the virus in Nigeria in 2017 and most notably during the 2022 global outbreak [4, 5]. Historically, MPXV infections within endemic regions have primarily been driven through zoonotic transmission with limited secondary human-to-human transmission and associated with close, extended contacts. These resultant mpox cases have been overrepresented among children <15 years in remote forested areas of Central and Western African regions [6]. In contrast, subclade IIb MPXV infections have been defined by sustained human-to-human transmission concentrated among adult males and within dense sexual networks. subclade IIb underwent rapid global expansion in 2022 and ongoing circulation among non-endemic regions[7].

Currently, an epidemiologic shift has been observed for Clade I MPXV in the Democratic Republic of the Congo (DRC). Reported mpox case numbers have risen at an alarming rate across historically endemic and non-endemic provinces within the country and across multiple risk groups within the population [8]. This led to the identification of subclade Ib and definition of subclade Ia [3, 9]. Early investigation of this outbreak has found that both subclade Ia and Ib are co-circulating [10] and currently, no specific transmission dynamics have been defined for either clade. The first cluster of subclade Ia mpox cases associated with sexual contact was identified in Kwango province in 2023 [11]. Subclade Ib, described in the Kamituga mining area, South Kivu province, is characterized by sustained human-to-human transmission and notable mutation within APOBEC3 target sites [3]. Subclade Ib subsequently spread to the highly connected and populated cities of Bukavu and Goma, North Kivu province, including internal displacement camps [12]. The shifting epidemiology of Clade I MPXV along with intra- and inter-continental exportations of cases (5) resulted in the declaration of the first Public Health Emergency of Continental Security by the Africa CDC and the second Public Health Emergency of International Concern (PHEIC) declaration for mpox by the World Health Organization [13, 14].

According to the DRC Ministry of Public Health, from 1 January to 30 September 2024, there have been 31,350 suspected cases of mpox reported across nearly all 26 provinces, with 992 deaths, resulting in a case fatality rate of 3.2%. The ongoing mpox public health emergency is the result of ongoing expansion of subclade Ib as well as multiple concurrent mpox outbreaks involving different subclade Ia variant lineages termed ‘groups’ (Group II, III, IV and V) [15]. While recent longitudinal phylogenomic analysis of subclade Ia genomes supports the notion that historic outbreaks of mpox in DRC have been driven predominantly by zoonotic transmission and multiple zoonotic introductions [15], observations from small outbreak clusters associated with these genomes have shown slight increases in APOBEC3 mutations suggesting that in some instances, more sustained human-to-human transmission may have been present.

Geographic expansion of Clade I MPXV since 2023 has included introduction to multiple large urban centers, including Kinshasa [10, 15]. Kinshasa is a diverse, densely populated, metropolitan area and migratory hub for both domestic and international travel. During the 2022 global outbreak, no rise in cases or introductions of subclade IIb were reported in the capital. However, with the increased reporting of secondary human-to-human mpox transmission, sustained mpox transmission in the most populous city on the African continent is of particular concern.

Here we sought to describe epidemiological trends of the escalating co-emerging subclade Ia and Ib mpox outbreaks in Kinshasa, DRC, focusing on confirmed cases identified during the ongoing public health emergency. Following the first PCR-confirmed case of mpox in Kinshasa, we report transmission pathways and demographic shifts in mpox clusters, with human-to-human transmission proposed as a primary driver of sustained urban outbreaks–an epidemiological trend far different from the rural, isolated cases characteristic of mpox cases in the past. Importantly, we further analyzed selected confirmed 2024 cases and characterized the genomic signatures of MPXV strains currently circulating in Kinshasa, including the introduction of APOBEC3-type consistent mutations in subclade Ia sequences. These observations increase concerns regarding further expansion of mpox due to more sustained human-to-human transmission in an urban center.

## METHODS

### St*udy design and population*

We conducted a retrospective observational study on suspected and confirmed mpox cases in Kinshasa, DRC. The study population included individuals with suspected and laboratory-confirmed mpox reported from January 2023 to September 26, 2024. Demographic data, laboratory results, and MPXV sequences were collected and analyzed to assess epidemiological trends, determine MPXV clade assignments, and perform phylogenomic characterization of MPXV variants that were circulating in Kinshasa.

Samples were collected as part of a routine countrywide mpox surveillance program and were exempt from informed consent procedures. Permission to use anonymized data from the mpox national surveillance activities for this report was granted by the Ethics Committee of the Kinshasa School of Public Health (ESP-UNIKIN, Number ESP/CE/05/2023).

### Sampling and Data Collection

Kinshasa, which is both a city and a province, is divided into 35 health zones. Mpox is a reportable disease in the DRC and is included in the nationwide Integrated Disease Surveillance and Response system [16]. As per national surveillance guideline, in response to a case alert, a health history and biological specimen is collected by a multidisciplinary team comprised of representatives from the Ministry of Health, the National Program for Mpox and Viral Hemorrhagic Fevers (PNLMPX-VHF), the Institut National de Santé Publique (INSP) and the Institut National de Recherche Biomédicale (INRB). Local surveillance teams collected data using the national case investigation form (Appendix Table 1), as well as samples from people with suspected mpox. This form includes information on demographic characteristics (age, sex, residence, including health zone and province, profession and nationality), time of onset of clinical symptoms, type of sample and sampling date of cases. As part of routine country-wide mpox surveillance, the INRB laboratory receives these samples from suspected mpox cases for molecular diagnosis and viral whole genome sequencing.

**Table 1.** Subclade, Age, and Sex of PCR-confirmed mpox cases in Kinshasa, DRC by period (n = 364)

| **Subclade** | **Total**<br>**N= 364** | **Period 1**<br>**Aug 2023 - Jun 2024**<br>**n=24** | **Period 2**<br>**July 2024-Sept 2024**<br>**n= 209** |
| --- | --- | --- | --- |
| la | 224 (61.5) | 24 (100) | 200 (58.8) |
| lb | 64 (17.6) | 0 (0) | 64 (18.8) |
| Unknown | 76 (20.9) | 0 (0) | 76 (22.3) |
| **Sex** |  |  |  |
| Male | 221 (60.7) | 17 (70.8) | 204 (60.0) |
| Female | 136 (37.4) | 7 (29.2) | 129 (37.9) |
| Unknown | 7 (1.9) | 0 (0) | 7 (2.1) |
| **Age** |  |  |  |
| Median (IQR) | 26 (19-32) | 23 (17-30) | 26 (19-33) |
| 0-5 | 21 (5.8) | 0 (0) | 21 (6.2) |
| 6-15 | 45 (12.4) | 5 (20.8) | 40 (11.8) |
| 16-25 | 109 (29.9) | 8 (33.3) | 101 (29.7) |
| 26-35 | 118 (32.4) | 9 (37.5) | 109 (32.1) |
| 36-49 | 65 (17.86) | 2 (8.3) | 62 (18.5) |
| Unknown | 6 (1.6) | 0 (0) | 6 (1.8) |

### Mpox Diagnosis and Whole Genome Sequencing

All samples (skin lesions (vesicles and crusts), oropharyngeal swabs, and blood) from people with suspected mpox were received and analyzed at INRB, Kinshasa. Viral DNA was extracted using the QIAamp DNA Mini Kit (Qiagen, Hilden, Germany) according to the manufacturer’s instructions. To confirm the diagnosis of mpox, a real-time PCR was performed with Orthopoxvirus- and MPXV-generic primers and probes, as described [17, 18]. Mpox-positive samples with a cycle threshold (Ct) value <31 underwent sequencing using either hybridization capture probe-based (Comprehensive Viral Research Panel, Twist Biosciences) or amplicon-based [19] enrichment before loading onto either the MiSeq or GridION sequencer. FASTQ files were processed using GeVarLi (https://forge.ird.fr/transvihmi/nfernandez/GeVarLi), CZ ID (https://czid.org/), Artic-mpxv-nf (https://github.com/artic-network/artic-mpxv-nf) or an in-house pipeline (Metatropics) pipeline for reference-based (GenBank accession number: NC_003310) consensus genome generation. Mpox-positive samples that remained without subclade assignment after sequencing were PCR-tested using a multiplex real-time PCR assay developed for MPXV detection and simultaneous subclade Ib identification (https://clinical.goldstandarddiagnostics.com/).

### Phylogenomic Analysis of MPXV genomes

Clade I MPXV reference genomes were obtained from NCBI GenBank, and Lusamaki et al. [15](accessed on 05/10/2024). We also included newly generated sequences from the ongoing national genomics surveillance (July-September: as available in https://github.com/inrb-labgenpath/DRC_MPXV_Genomic_Surveillance). Duplicate sequences were removed, and the dataset was filtered to include MPXV genomes meeting specific quality criteria: over 80% horizontal coverage (fewer than 20% ambiguous bases), exclusion of sequences flagged by IQ-TREE [20], and complete metadata for collection date and location. This dataset was then combined with consensus genomes from Clade I mpox cases in Kinshasa, DRC, using the same quality thresholds. The subclade IIa MPXV sequence (accession number: AY603973) was included as an outgroup. The entire MPXV genome dataset was aligned to the Clade I reference (accession number: NC_003310) using Squirrel (https://github.com/aineniamh/squirrel), with low-complexity and repetitive regions masked from the alignment. A maximum-likelihood phylogenetic tree was inferred using IQ-TREE2 v2.1.4 [20], with the ‘K3Pu+F+I+R5’ substitution model selected as the best fit. Branch support was determined through ultrafast bootstrap approximation with 10,000 replicates [21].

We then focused on the established Kinshasa Clade I MPXV outbreak genomes and reconstructed individual mutations onto each branch of the maximum likelihood tree, using ancestral state reconstruction in IQ-TREE2 v2.1.4 [20]. Squirrel then assigns these mutations into two types: non-APOBEC3-type mutations, caused by errors due to the polymerase at replication, and APOBEC3-type mutations, identified based on their dinucleotide context and specific transitions [22].

## RESULTS

As part of routine countrywide mpox surveillance, the INRB laboratory received 1024 samples from suspected mpox cases in Kinshasa, reported between January 2023 and September 26th, 2024. Of these, 364 (35.6%) were PCR-positive for MPXV; the first case was sampled on August 18, 2023. Between August 18, 2023 and June 30, 2024, 218 mpox cases were reported for investigation-24 (11%) of these were laboratory confirmed cases and all were identified as subclade Ia. The first confirmed subclade Ib mpox case in Kinshasa was reported on July 1, 2024. Between July 1 and September 26, 2024 806 cases were reported of which 340 were mpox confirmed (42%), 58.8% and 18.8% identified as subclade Ia and Ib, respectively, 22.3% were unknown (Figure 1).

**Figure 1.**
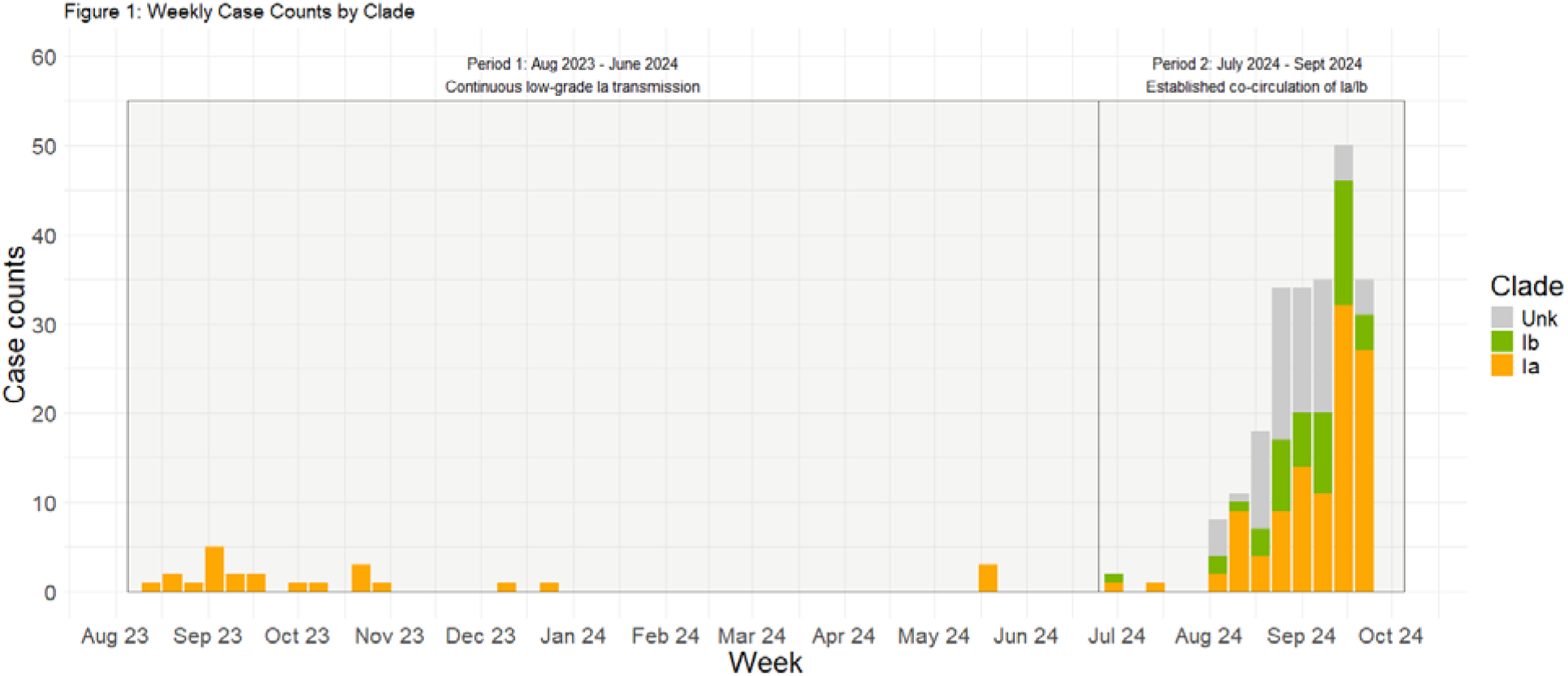
Epidemiologic curve of PCR-confirmed Kinshasa mpox cases (August 2023-September 2024, n = 364)

Among mpox cases, 221/364 (60.7%) were male, with median age 26 years (interquartile range 19-32) (Table 1). Children 15 years or younger comprised 18.1% (66/362) of confirmed cases, individuals aged 16–49 years accounted for 80.2% (292/364). Most confirmed cases were male; 54.2% of cases under 15 years of age were male, 62.3% of cases over 15 years old were male.

This epidemiologic curve suggests two distinct periods in the Kinshasa 2024 outbreaks. An initial period from August 2023 to June 2024 characterized by sporadic subclade Ia mpox case detections, and a second period from July 2024-October 2024 characterized by co-emergence of both subclade Ia and Ib. The spatial, sex and age-group distribution of confirmed mpox cases by subclade in Kinshasa for these periods is shown in figure 2. Of note, Limete Health Zone (HZ) reported the most confirmed mpox cases over the course of Period 2 – of both subclades (Figure 2c).

**Figure 2.**
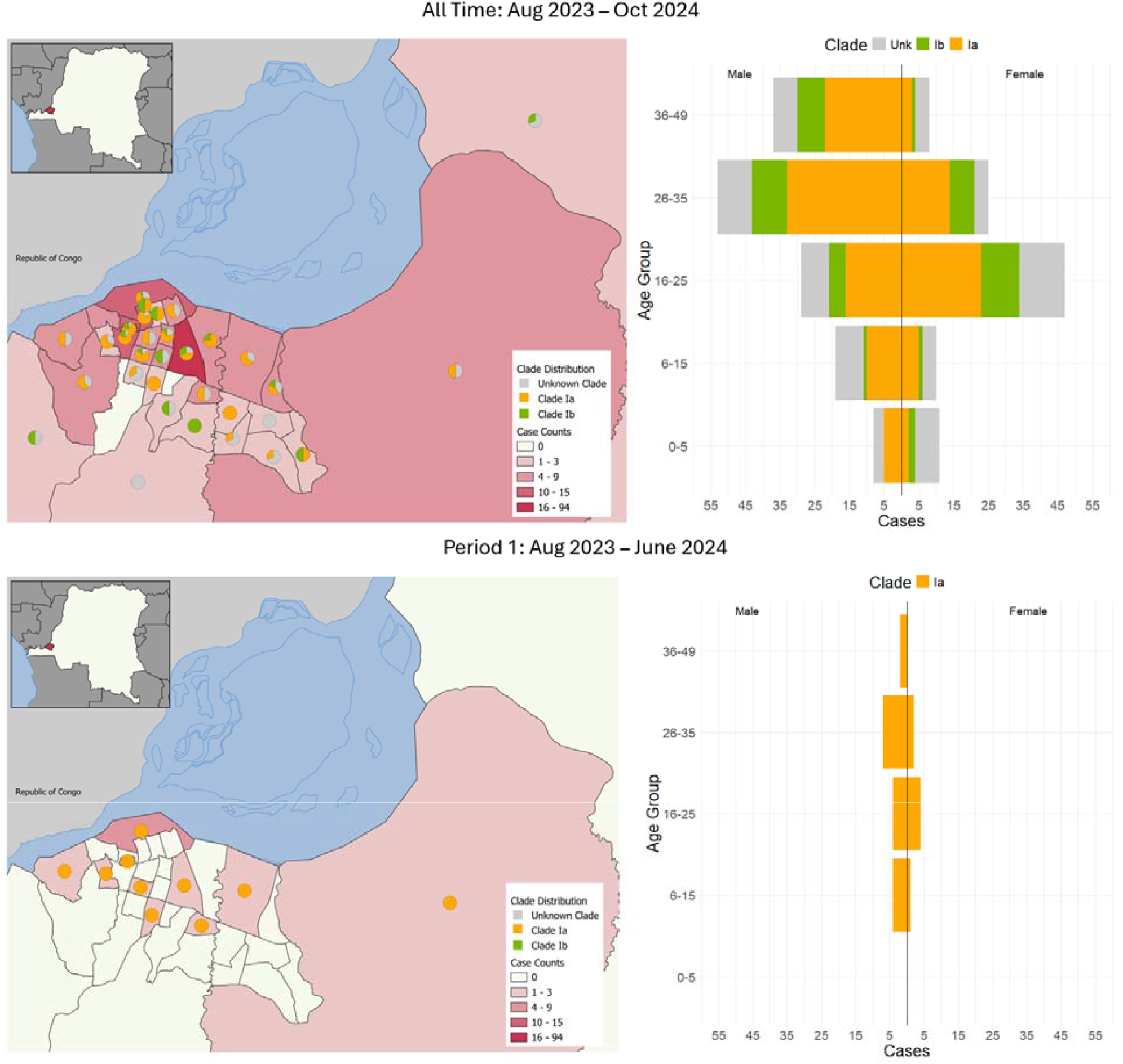
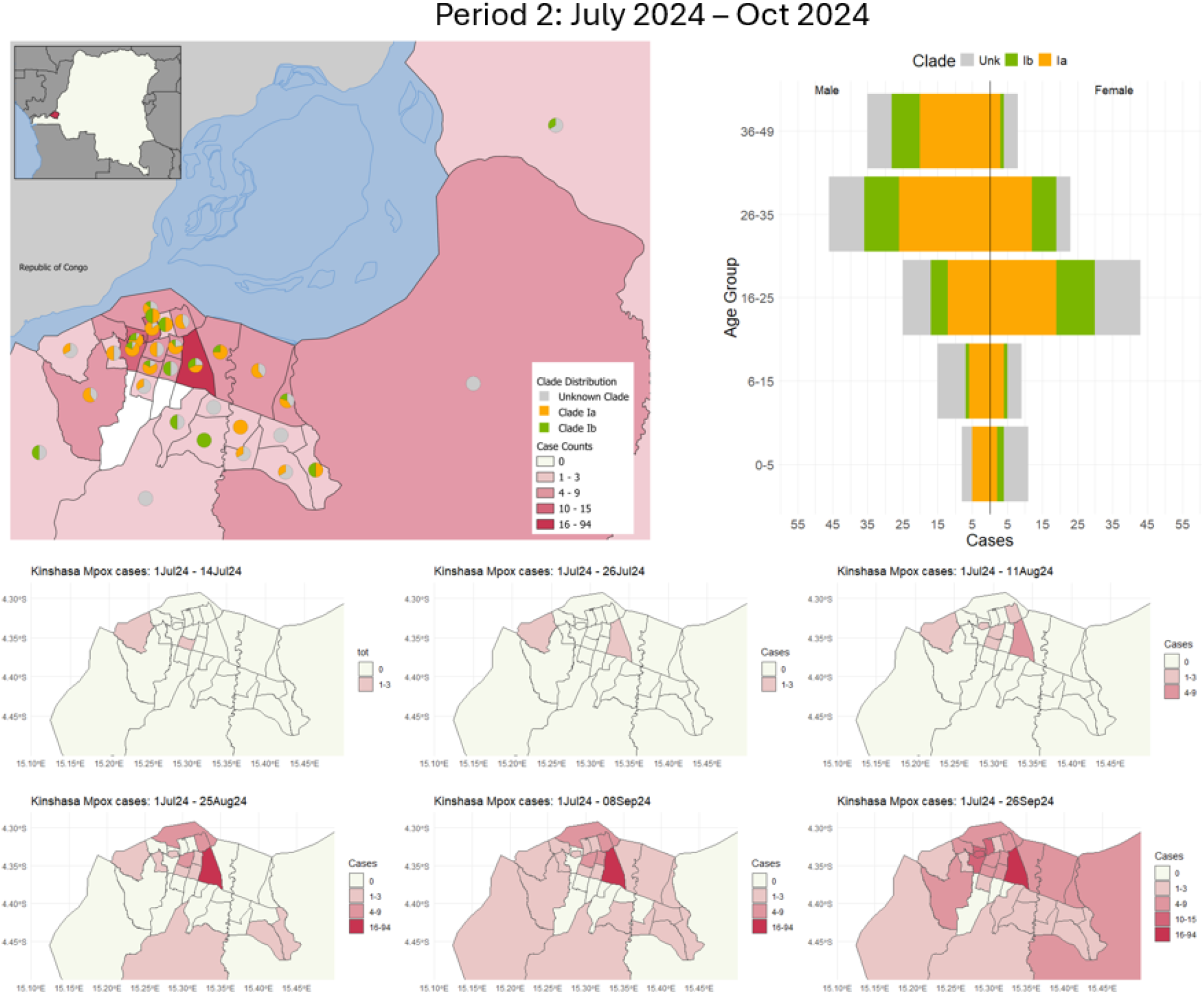
Mpox cases in Kinshasa by age, sex, and subclade over time. Left: Spatial distribution of confirmed mpox cases by subclade in Kinshasa by health zone. Right: distribution of confirmed mpox cases by age group, sex, and subclade. Panel a: total study period August 2023-September 2024, Panel b: period 1 August 2023-June 2024, Panel c: period 2 July 2024-September 26, 2024.

### Phylogenomic Analysis of Mpox-positive Samples

All MPXV genomes (n=11) preceding the local-established mpox outbreaks belong to 5 clusters, (over the period August 2023-June 2024) belong to subclade Ia Group II, and do not cluster with the local-established mpox outbreak in the period July 2024-October 2024, despite also belonging to the subclade Ia Group II. This suggests that the sporadic Ia cases observed in August 2023-June 2024 resulted from independent introductions and a separate single introduction has led to the June 2024-September 2024 subclade Ia outbreak (Figure 3). The closest neighbors of the subclade Ia outbreak cluster are genomes identified in mpox samples from the Republic of Congo (Cuvette-Central) and the Equateur Province, DRC, and shows links with more recent genomes from Kwilu, Kwango, and Kongo-Central. The subclade Ib outbreak, however, seems closely interlinked with other DRC Provinces, such as South Kivu, as well as Kasaï, Tanganyika and Tshopo. 63% Of mutations in subclade Ia (Figure 4) and 66% of mutations in subclade Ib (Figure 5) are consistent with APOBEC3-driven changes, a hallmark of human-to-human transmission [22].

**Figure 3.**
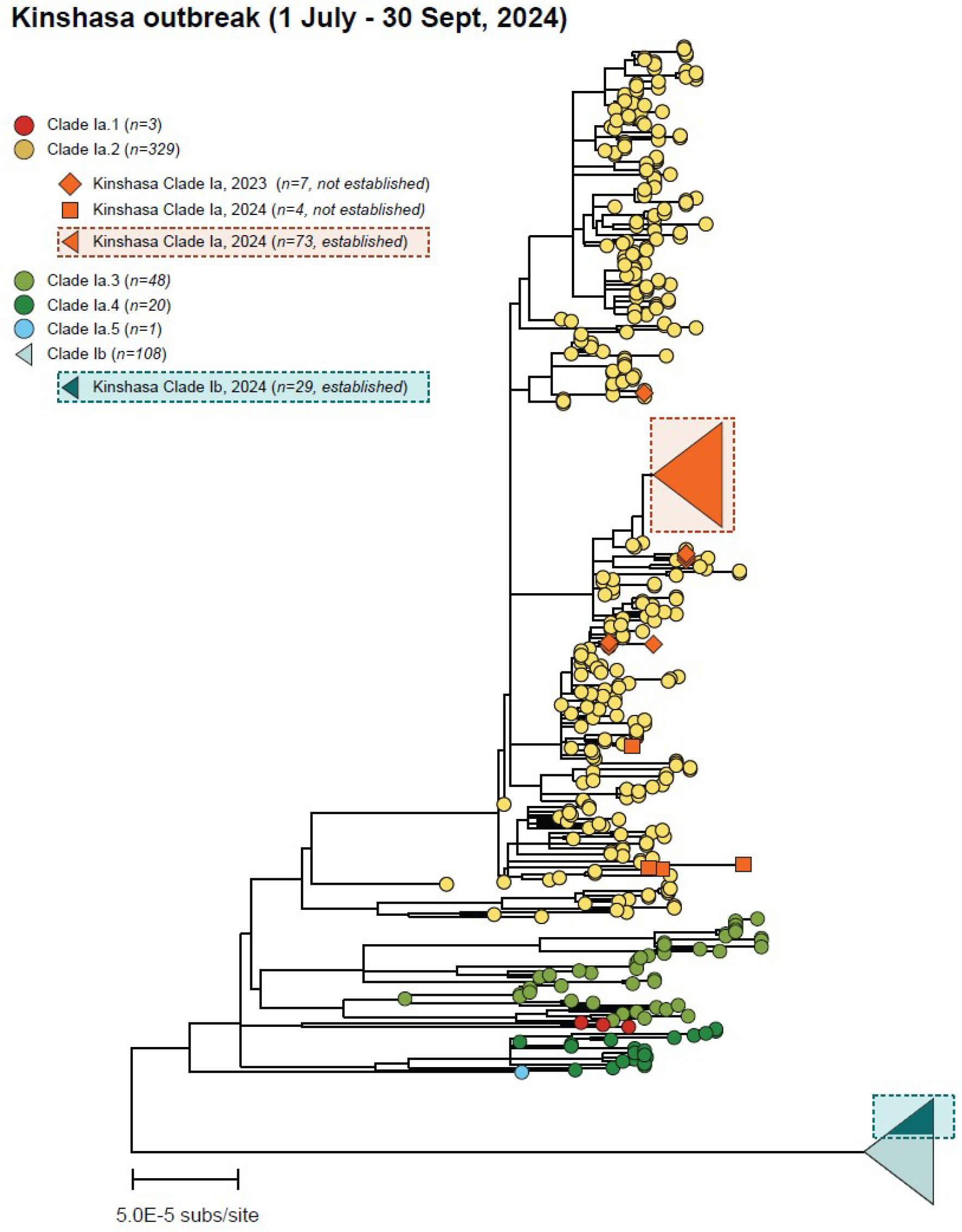
Maximum likelihood phylogeny: Subclade Ia and Ib MPXV have reached Kinshasa, the capital of the DRC, to cause two co-emerging local outbreaks. The maximum likelihood phylogeny shows that viral genomes isolated from mpox patients residing in Kinshasa, in 2023-2024 belong to subclade Ia (Orange, total n=93; established n=73; not established n=11) and Ib (Green, established n=29). Non-established Kinshasa subclade Ia observations are indicated with orange diamonds (2023) and squares (2024). Subclade Ia ‘groups’ are indicated as: I (red), II (yellow), III (light green), IV (dark green), V (blue). The (established) Kinshasa Ia and Ib outbreaks are marked with rectangles with dashed borders.

**Figure 4.**
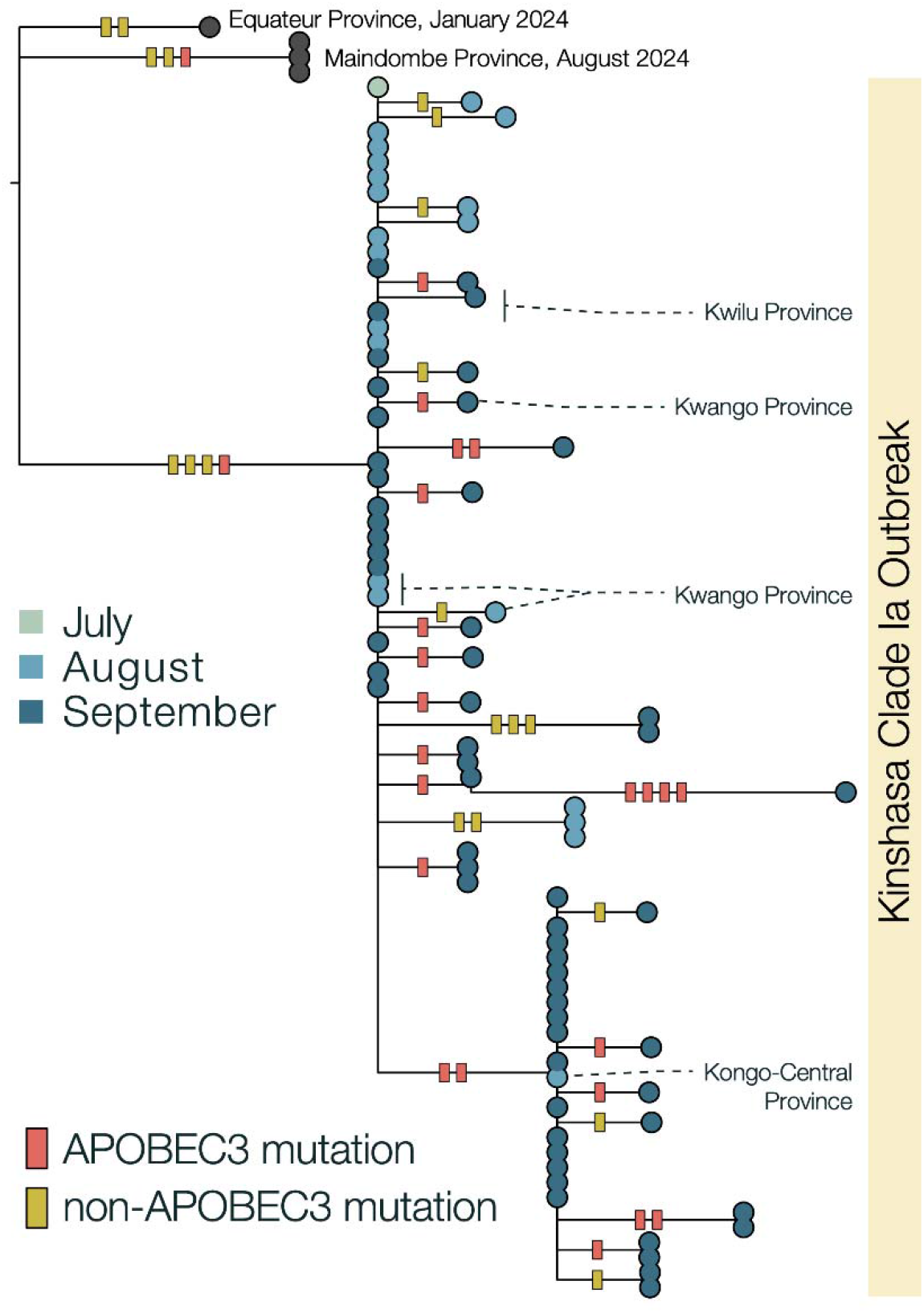
Maximum likelihood tree and APOBEC3 reconstruction of a Kinshasa 2024 outbreak of MPXV subclade Ia: 63% (22/35) of the mutations in the ongoing 2024 Kinshasa subclade Ia outbreak, indicative of APOBEC3-induced changes in sustained human-to-human transmission. The subclade Ia Kinshasa outbreak genomes are shown annotated by month of sample collection in 2024. APOBEC3 driven changes are indicated by red bars on the branches, others are indicated as yellow bars. Outgroup genomes are indicated by black dots.

**Figure 5.**
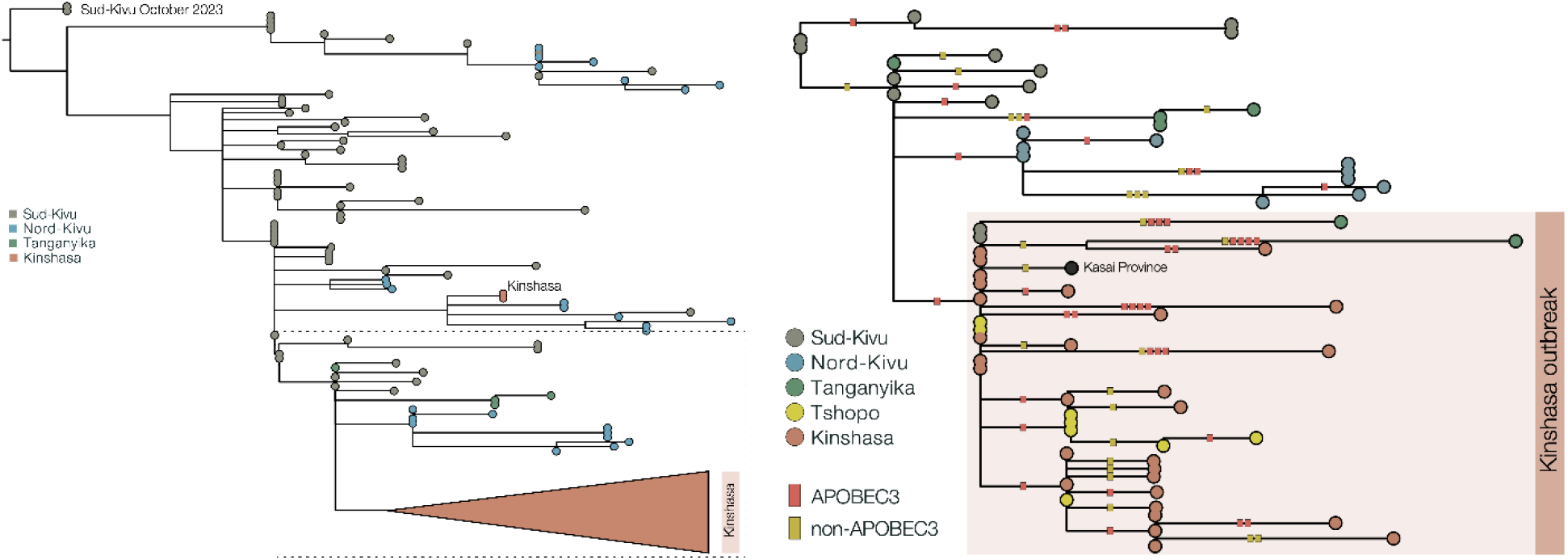
Maximum likelihood phylogeny of country-wide subclade Ib outbreak (top panel) and APOBEC3 analysis of subclade Ib, Kinshasa 2024 outbreak (bottom panel): 66% (29/44) of the mutations in the ongoing 2024 Kinshasa subclade Ib outbreak (within the box) are consistent with APOBEC3-driven changes. This is consistent with the 60% (88/146) mutation fraction observed for the Ib outbreak overall (not shown). 2024 Kinshasa subclade Ib genomes are shown annotated by province (Sud-Kivu, Nord-Kivu, Tanganyika, Tshopo, and Kinshasa, with one additional genome from Kasaï Province). APOBEC3 driven changes are indicated by red bars on the branches, others are indicated as yellow bars. We annotated using the province instead of month of sample collection to highlight the geographic interconnectivity of the Kinshasa subclade Ib outbreak, especially with Tshopo province.

## DISCUSSION

Here, we report continued transmission chains and repeated introduction of both MPXV subclades Ia and Ib in Kinshasa, DRC. This evidence provides further support that mpox cases in Kinshasa have been related to multiple concurrent clusters over the past two years rather than a singular outbreak. By separating reported clusters into two “periods” this may indicate an important shift in mpox transmission: Period 1 was characterized by the first confirmed subclade Ia MPXV case in Kinshasa followed by localized spread and limited introduction from endemic provinces (e.g. Mai-Ndombe), whereas Period 2 was indicated larger, sustained outbreaks of both subclades, which may be linked to increased human-human transmission.

Prior to July 2024, the distinct transmission chains in Kinshasa associated with Period 1 indicate an R_0_ of less than 1. Demographic data for Period 1 presents almost equal distribution of cases among children – male and female – which is more indicative of household transmission. However, the rapid increase in cases during Period 2 suggests that other factors are contributing to maintain mpox transmission. While transmission via sexual networks has been suggested as the primary driver of mpox expansion in urban centers like Kamituga [3], the contributions of nosocomial transmissions or in households cannot be dismissed, which have been linked to the Period 1 clusters.

Beyond routine surveillance data, in-depth case investigations identified three small, self-limiting clusters in August and September 2023. Cluster 1 originated from an adult male traveling via boat to Kinshasa from Inongo, Mai-Ndombe. Case investigation and contact tracing found that mpox had been transmitted from this index case to four primary contacts–a coworker, two healthcare workers, and 1 nosocomial contact–and then to three household contacts-of-contacts. During this same period, two additional 3-person clusters were identified, transmission was determined to be via household and social contact. Samples collected from these clusters were later identified as subclade Ia MPXV-positive. This investigation into these early cases of mpox demonstrates the endemicity of MPXV in the DRC– multiple concurrent introductions can be observed among multiple populations, age and sex groups, through multiple transmission pathways. However, with the increased reporting of mpox in Period 2, reciprocal case investigations were not completed for all confirmed cases – therefore, it is impossible to definitively confirm the mode of MPXV transmission.

This confirmation of human-to-human transmission in Kinshasa has major implications for both mpox containment and mitigation in the DRC as well as for broader international expansion. As a major transit hub and the second largest city in Africa with ∼17 million inhabitants (https://www.statista.com/statistics/1218259/largest-cities-in-africa/), Kinshasa is a prime location for rapid viral spread and evolution. Expanding upon the first reported co-circulation of subclades Ia and Ib [10], we present confirmed human-to-human transmission chains of subclade Ia MPXV in 2023, as well as continued co-emergence of both subclades in the capital.

Furthermore, Period 2 epidemiologic data also suggests that subclades Ia and Ib are not as epidemiologically distinct as previously reported - we report confirmed cases of both subclades among majority adolescents and adults with probable sexual contact transmission, a shift from the historic cases reported among children in rural environments [6]. Specifically, during Period 2, we observed the most cases irrespective of subclade among women 16-25 and men 26-35 years old; this demographic presentation is characteristic of transmission through sexual contact. Spatiotemporal analysis of reported cases highlight Limete HZ as a center of mpox transmission – this HZ is known to have a higher density of professional sex workers, which may also be playing a role driving transmission. With the recent deployment of the JYNNEOS vaccine in DRC [23], these profiles may help to better define which populations should be considered “at-risk”.

The epidemiological profiles of those PCR-positive cases underscore the value of regular mpox surveillance as the definition of “at-risk” populations continues to evolve. However, the epidemiological data from Kinshasa, and more broadly in DRC, has limitations. While some suspect cases undergo in-depth investigation by teams at the INRB and PNLMPX-VHF, most are surveyed and sampled as part of routine surveillance with PCR-testing conducted at the INRB – investigation forms may also be completed by hand yet, are not always readily accessible. Furthermore, laboratory testing is only paired with limited epidemiological data - such as age and sex - and does not always include invaluable information such as demographic characteristics, risk factors or clinical presentations. Similarly, suspect cases are not always followed longitudinally, making it difficult to link reported cases with case fatalities. Data missingness, and with this, the lack of well-defined transmission chains, are key factors which may provide additional insights into the two subclades if further investigated. Additionally, the mpox profiles presented here are products of the Kinshasa sampling strategy - which may not capture the entire outbreak. As these outbreaks have been described in a nonendemic region of the country, mpox identification and sampling in Kinshasa may not capture all cases.

Due to the high fidelity and error correction of the MPXV DNA polymerase, mutations during genome replication occur at a much lower rate than those induced by APOBEC3 (∼1 nucleotide change every 3 years vs ∼6 per year respectively) [22, 24]. Thus, over a short timescale during human-to-human transmission, we expect APOBEC3-type mutations to predominate and occur at a sufficient rate to reconstruct the patterns of transmission and spread of the virus. In this analysis, we estimate that 63% of mutations in subclade Ia and 66% of mutations in subclade Ib are consistent with APOBEC3-driven changes, a hallmark of human-to-human transmission [22]. Although sequencing can introduce errors that manifest as artifactual mutations, these are unlikely to resemble the specific characteristics of those induced by APOBEC3. A small fraction of polymerase-induced mutations may, by chance, appear similar to APOBEC3-induced mutations (∼ 8% is estimated for Clade I) [22], but their absolute rate is negligible over short timelines. Given the predominance of APOBEC3-type mutations in these outbreaks, specifically in the subclade Ia outbreak in Kinshasa and the entire subclade Ib outbreak first observed in South Kivu province [3, 9], we can be confident that they both resulted from sustained human-to-human transmission (following a single zoonotic introduction). This represents a shift in the epidemiology of subclade Ia group outbreaks, which had been suggested to be driven by multiple independent zoonotic introductions, until now. This outbreak should, therefore, be closely monitored, as it represents a significant threat for regional and international dissemination as illustrated with new sequences from Kwilu, Kwango, and Kongo-Central (subclade Ia), as well as Kasaï, Tanganyika and Tshopo (subclade Ib), linked to Kinshasa genomes. The potential link with sexual networks should be further examined.

In pairing epidemiological surveillance data with genomic analysis, we suggest increasing human-to-human transmission of both subclades of MPXV Clade I among the greater Kinshasa population.

## Data Availability

All data produced in the present study are available upon reasonable request to the authors

